# The Effects of Long COVID-19, its Severity, and the Need for Immediate Attention: Analysis of Clinical Trials and Twitter data

**DOI:** 10.1101/2022.09.13.22279833

**Authors:** Arinjita Bhattacharyya, Anand Seth, Shesh Rai

## Abstract

The coronavirus disease 2019 (COVID-19) has been declared a pandemic since March 2020 by the World Health Organisation (WHO). The infection pathway follows symptoms of fever, cough, shortness of breath, dyspnea, and severe cases that lead to hospitalization, emergency life support, and even death. Identifying the disease progression and predicting patient outcomes early, precisely predicting the possibility of long-term adverse events through effective modeling, and use of real-world data such as longitudinal clinical trial data, electronic health records data, and health insurance data are of immense importance to effective treatment, resource allocation, and prevention of severe adverse events (SAE) of grades four or five.

The main goal of the study is threefold. Firstly, we raise awareness about the different clinical trials that are being conducted concurrently on Long covid-19, and how these are beneficial. Secondly, we analyze the recent tweets on Long haul covid-19 and give an overview of the sentiments of the opinion of the people. Finally, we analyze the sentiment scores and find if they are associated with the demographics of the tweeters via a negative binomial regression model.

The trials were selected with long Covid-19 available in ClinicalTrials.Gov. Also, the tweets obtained with the term #long covid-19 consisted of 8436 tweets. We utilized the NRC Emotion Lexicon method for sentiment analysis is a list of English words and their associations with eight basic emotions (anger, fear, anticipation, trust, surprise, sadness, joy, and disgust) and two sentiments (negative and positive) (11). We obtained a matrix of sentiment scores, as well as retweet counts and favorite counts which were analyzed. We regressed the retweet counts and the favorite counts with the sentiment scores and find if they are associated with the emotions and sentiments of the tweeters via a negative binomial regression model since the outcome variable is count data.

Our results find that there are two types of clinical trials (a total of 298) being conducted 1)observational and b) interventional. The details about enrollment, time to completion, clinical trial phases, etc., are discussed. Sentiment analysis with the NRC method of the tweets shows that there are both positive and negative sentiments. The retweet counts and favorite counts are associated with the sentiments and emotions such as disgust, joy, sadness, surprise, trust, negative, positive, etc.

Finally, to conclude we need resources, and further research needs to be conducted in this area of long Covid-19.

## Background

The coronavirus disease 2019 (COVID-19) has been declared a pandemic since March 2020 by the World Health Organisation (WHO). There have been 426,551,362 cases and 5,898,442 deaths worldwide since its advent in December 2019. The SARS-Cov-2 virus has been transmitted to humans and has produced several severe, deadly, and highly infectious variants through mutations. The infection pathway follows symptoms of fever, cough, shortness of breath, dyspnea, and severe cases that lead to hospitalization, emergency life support, and even death. Identifying the disease progression and predicting patient outcomes early, precisely predicting the possibility of long-term adverse events through effective modeling, and use of real-world data such as longitudinal clinical trial data, electronic health records data, and health insurance data are of immense importance to effective treatment, resource allocation, and prevention of severe adverse events (SAE) of grades four or five. There are significant research needs for such situations, thus helping to allocate the ‘right drug to the right patient.’ The primary goals of this study are to address the current initiatives through clinical trials of the Covid-19 infection, and the prevalence of long-haul Covid-19, and to raise the need for prediction of the immediate and long-term outcome of Covid-19 through a novel prediction algorithm. There is also a need to assess the importance of demographics, genetic biomarkers, comorbidities, concomitant medication, social and genetic or environmental, or economic effects having a specific role in predicting the disease’s outcome. Further conducting subgroup findings through a novel methodology will help to stratify patients having similar results that will help us identify treatment allocation strategies.

To support this future research may use survival and binary logistic regression models depending on the outcome of interest. Further, depending on the availability of data, research can be done to create novel methodology on variable selection and shrinkage prior methods of Lasso, Elastic-Net, and Bayesian shrinkage estimation to find the most significant variables responsible for such a long Covid-19 effect. Finally, whether the person will develop long-term covid-19 can be predicted. The groups of patients with similar outcomes need to be stratified and treatment interventions nneedto be allocated accordingly. The novel techniques need validation by simulations, and applied to real-world EHR, and clinic al trial data.

We anticipate finding that age, sex, gender, race, vaccination status, concomitant medications, prior illness, comorbidities, and symptoms may be responsible factors contributing to the disease’s progression. The short-term outcomes may depend on these factors, along with the other covariates such as recovery, time varied symptoms, genetic factors, interaction effects, hospitalization, and ventilation status, as some of the responsible factors.

Based on the accuracy of the methods, the long-term presence of SARS-CoV-2 RNA in humans and its dependency on demographics, genetics, and interaction effects need to be addressed, the potential long-term consequences for Covid-19 survivors will impact patients and need public health, government, and health policy makers’ further attention to mitigate such concerns and will encourage the development of therapeutics, This research work lays the stepping stone for future guidance and reports the currently available clinical trials and describes them. It also lays the ground for understanding public opinion toward Covid-19 by analyzing tweets from people on the social media

## Motivation

In late December 2019, a series of pneumonia cases of unknown origin emerged in Wuhan, China, with clinical presentations resembling viral pneumonia. Deep sequencing analysis of lower respiratory tract samples indicated a novel coronavirus named 2019 novel coronavirus (2019-nCoV), thus creating a worldwide concern (1). Two years ago, this ‘severe acute respiratory syndrome coronavirus 2’ (SARS-CoV-2) virus was declared a global pandemic. As we write this proposal, COVID-19, officially named by World Health Organization (WHO), has a case count of 426,551,362 cases and 5,898,442 deaths worldwide (as of Feb 2022), according to John Hopkins’s Coronavirus Resource Center (2). The COVID −19 pandemic is the most significant global crisis since World War II that affected almost all the countries on Earth, according to the United Nations (3).

As of May, 27, 2022, there are 7695 studies listed concentrating on Covid-19, of which only 19 are dedicated to understanding the long-term effects of Covid-19. Out of the 19 studies, one is reported as available, three are completed, one is enrolled by invitation, three are not yet recruiting, 1 study has been withdrawn, and ten are currently recruiting. Only one concluded study has results (NCT04871815, “Effects of Sodium Pyruvate Nasal Spray in COVID-19 Long Haulers). Eight of them are treatment by administration of drugs, and one is device-related. The outcome measures include serious adverse events (SAE), short-term and long-term feature changes, and demarcation between symptoms of severe vs non-severe cases, among others. All the studies included all genders except one study that only enrolled the female population (NCT05225220, “Multimodal Investigation of Post COVID-19 in Females”) with an intervention of the device. The sponsors included both universities and hospitals. The median age group was 18 years or older (65yrs or more). One of the studies (NCT04956445, “Collection of SARS CoV-2 (COVID-19) Virus Secretions and Serum for Countermeasure Development”) included a child of ages six months or older. Most of the studies were either interventional (63%) or observational (31%). Six studies reached the phase 2 stage of the clinical trial. 31% of the studies are industry-sponsored. The maximum enrollment was 2000, with 20 as the least number of participants enrolled. Seven study designs are allocated randomly. The earliest start date of a trial is as early as March 17, 2020, with the recent study being on May 3, 2022. Brain Fog is one such severe condition, and the neurological effect of exposure to this virus is of grave concern. Only one study focuses on this post-trauma and neurological aspect of Covid-19 (NCT5042466). This necessitates attention towards research needs in the covid-19 with extended effects where biostatisticians have a lot to contribute.

## Existing Research

Since the pandemic, there have been many articles; 452 publications existed on Covid-19 from November 2019-to May 2020 (4), most of which do not focus on long-term covid infections. Studies have shown that long-haul Covid-19 is prevalent in 25% (1 in 4) of Covid-19 patients, irrespective of severity. Covid-19 patients, after recovery, continue to suffer for weeks and months. Research can help answer the questions on the long-term consequences of the disease from our patients and healthcare providers. A recent meta-analysis study on the global prevalence of Post-Acute Sequelae of COVID-19 (PASC) or Long COVID showed that there was a global long covid-19 prevalence estimate of 0.43 (95% confidence interval [CI]: 0.35, 0.63), with females having a high rate of prevalence than males, with regional prevalence having highest in the Asian population followed by Europe and US. Among commonly reported PASC symptoms, fatigue and dyspnea were reported most frequently, with a prevalence of 0.23 (95% CI: 0.13, 0.38) and 0.13 (95% CI: 0.09, 0.19), respectively. The global pooled PASC prevalence decreased for 30 to 90 days after the index test positive date (5).

Although it is anticipated that most of the COVID-19 symptoms disappear within 21 days, nearly 10% of COVID-19 patients show signs even after three weeks, 5% for eight weeks or more, and 2% suffer for almost three months. The long-haul patients after recovering from severe symptoms of shortness of breath, chest pain, most common-brain fog, fever, and headache, among others, irrespective of the variant of infection, and age with 20-30% of the children having long Covid symptoms. There are concerns about permanent damage to the lungs due to the covid-19-related symptoms of acute respiratory distress syndrome (ARDS) and Alzheimer’s disease due to the most common brain fog, pulmonary, cardiovascular, neurological effects, and unknown inflammation. To understand this newly developed disease, further research, attention to the needs of the patients through follow-ups, and the creation of post covid clinics are essential.

Some notable research has studied the impact of nutrition and high-fat diets that lead to diabetes and obesity, contributing to the long-term effects of Covid-19 [6]. There were about more than 50 long-term symptoms reported in a systematic review, of which the most common are fatigue, headache, attention disorder, hair loss, and dyspnea [7].

Case reports have also suggested that children may experience similar long Covid symptoms, of which females may be more affected [8]. One of the significant concerns of long-haul COVID-19 is that the patient is unaware of the long-term effects, which cannot be predicted at the early stage of the disease. Also, both adult and pediatric populations are affected by it. The participation of an international and interdisciplinary group of researchers with the availability of big data such as electronic health record (EHR) data set, clinical trial data, and medical insurance data are essential. Such data in a combination of novel methodologies will help understand the consequences and characteristics of both local and global Covid-19 affected populations. This universal view from research studies will be helpful in international health policy, opening Covid-19 long-term clinics and health insurance policies [5]. There has been ML and standard statistical models developed for task but we need to understand the data and address such needs, however, the targeted population, and inference derived from such should be cautiously interpreted. Often these models condition on patients testing positive for COVID-19, but that may introduce bias depending on who has access to testing/care. Choosing the right control group is key to obtaining a valid prediction model. Each prediction model has its pros and cons and assumptions that must be justified (10).

## Limitations & Research Questions

The collection of EHR and clinical trial data on both adult and pediatric populations is essential to address the following research questions.The research topics and questions that can be addressed are as follows:

What is the prevalence of the long-term effects on Covid-19 patients who have recovered from the disease?

What are the symptoms of long-haul Covid-19? What are the demographics of long-haul Covid-19 patients?

What factors other than the Covid-19 infection include demographics (sex, age, race), vaccination status, concomitant therapy, hospitalization, life support or intensive care unit, comorbidities that influence long-term Covid-19?

How are the long-term Covid-19 treated, and what are the treatment and resources allocated for such disease? How do they vary according to symptoms?

Are there any genetic, gene-treatment, or gene-environment factors responsible for such a disease? Brain fog is one of the long-term symptoms, and there are growing concerns about neurological disease progressions as a long-term consequence of Covid-19. The effect of such a claim needs to be validated by data that can be achieved through this research.

Also, we are interested in the long-term effects and mental health aspects of the first responders, such as physicians, nurses, and hospital authorities.

Were patients recovering from intensive care or life support due to hospitalization or having comorbidities more prone to long-haul Covid-19 symptoms than those who recovered at home? Does a healthy lifestyle influencepositively towards long term outcomes of covid-19?

What are the long-term effects and treatment in the pediatric population?

Is the prevalence of long-term effects similar to previous infections such as MERS and influenza? How does the mortality of Covid-19 vary across symptoms, demographics, comorbidities, and regions?

Identify the subgroups of adult and pediatric patients that are more prone to the long-term effects of covid-19 and mortality.

Whether anti-virals and current medications be utilized for potential benefit to such long haulers of covid-19?

Whether there are drug-drug interactions lead to long-term effects on covid-19 patients with comorbidities?

What precautions can protect against long-term effects, does vaccination and booster dose, wearing masks help?

Is there a dominant variant that leads to long covid-19 symptoms?

## Methodology

The main goal of the study is threefold. Firstly, we raise awareness about the different clinical trials that are being conducted concurrently on Long covid-19, and how these are beneficial The trials are available on ClinicalTrials.Gov and chosen with the search term “long Covid-19” (Figure 1 a) We mostly analyzed this data through effective data visualization techniques.

**Figure 1a.**
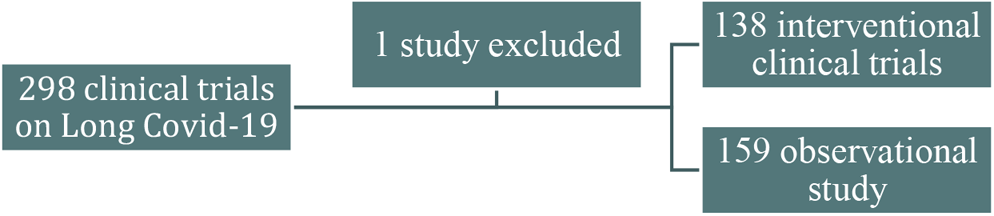
The Clinical Trial Studies included in the research work Secondly, we analyzed the recent tweets on Long haul covid-19 and give an overview of the sentiments of the people’s opinion.

One way to analyze the sentiment of a text is to consider the text as a combination of its individual words and the sentiment content of the whole text as the sum of the sentiment content of the individual words. We utilized the “NRC Emotion Lexicon” method for sentiment analysis is a list of English words and their associations with eight basic emotions (anger, fear, anticipation, trust, surprise, sadness, joy, and disgust) and two sentiments (negative and positive) (11). We obtained a matrix of sentiment scores, as well as retweet counts and favorite counts which were analyzed. We regressed the retweet counts and the favorite counts with the sentiment scores and find if they are associated with the emotions and sentiments of the tweeters via a negative binomial regression model since the outcome variable is count data.

### Reporting results on Long Covid-19 from clinicaltrials.gov

As of Sep 5, 2022, there are 297 studies posted on CliniTrials.Gov where research is concentrated towards PASC or Covid-19. The status of the studies is shown in the pie diagram in Figure 1b.

**Figure 1b:**
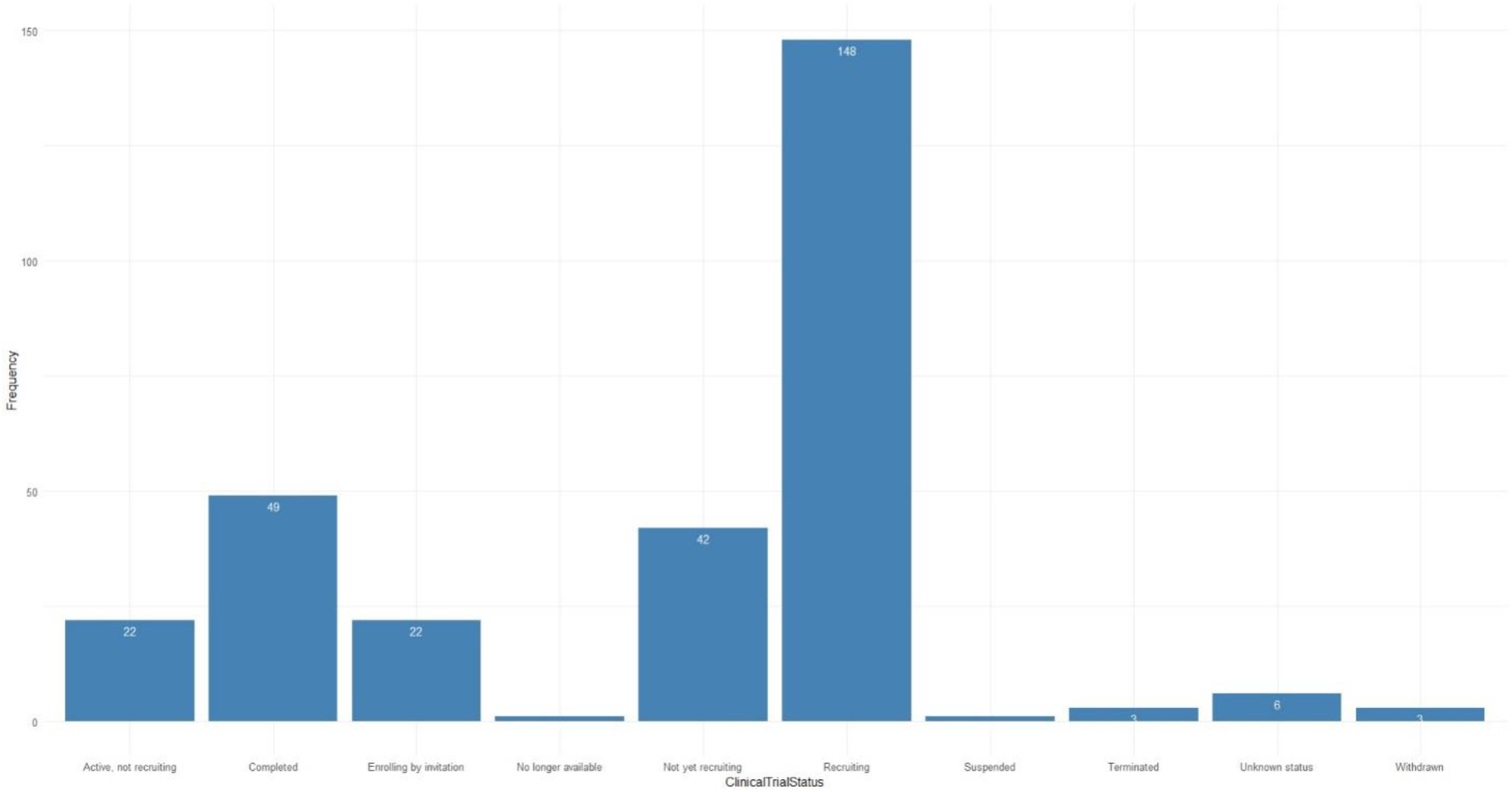
Status of Clinical Trials for Long- Covid-19

There are 294 trials where both males and females can be enrolled, two trials had a restriction for males, and one had a restriction for females. Figure 2 represents the word cloud of all the long-covid-19 trials. Figure 3 shows the word clouds for a) titles of interventional trials b) interventions in the trials and c) outcome measures of interventional trials

**Figure 2:**
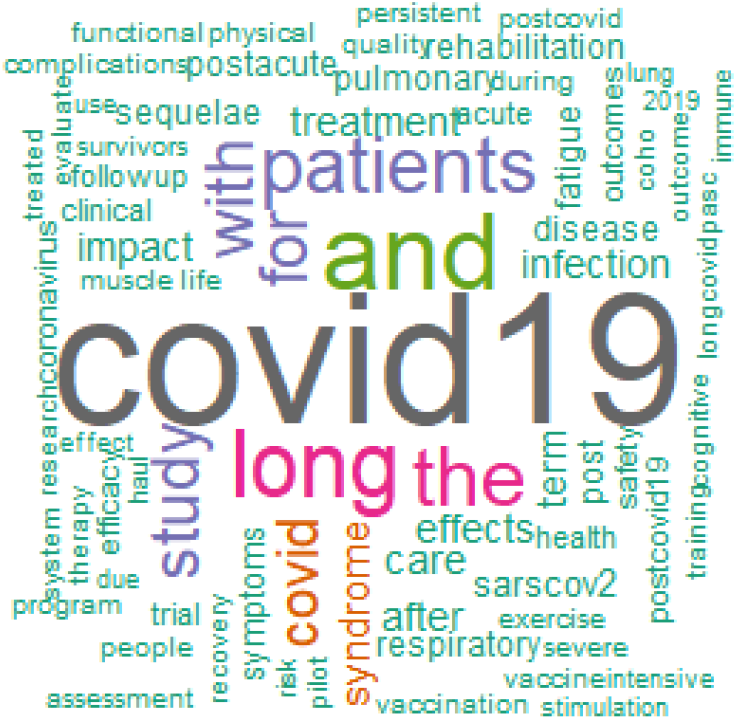
Wordcloud of Titles of all the Long- Covid-19 Trials

**Figure 3:**
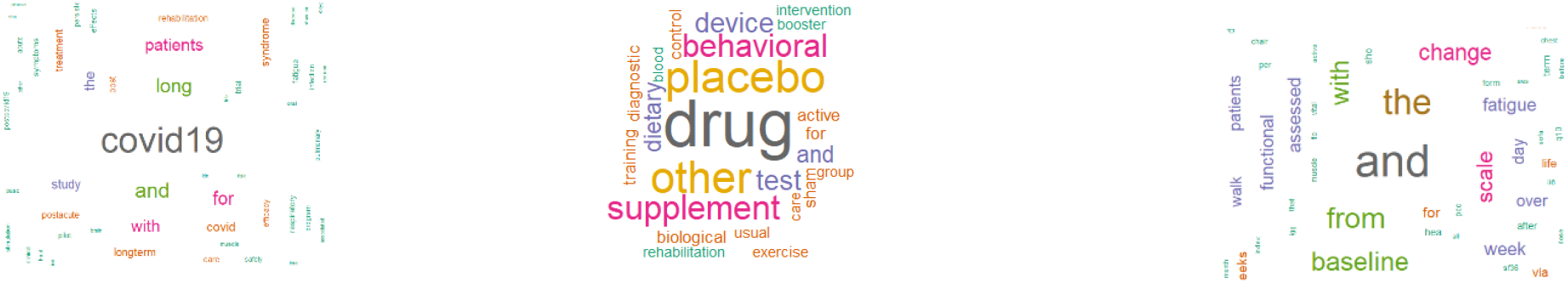
Wordclouds a) Title of Interventional Trials b) Intervention in the Trials c) Outcome measures of Interventional Trials

There are 159 trials that are observational or other types. We have discarded the trial “NCT04798066” while analyzing the observational studies.

There are 138 trials that are interventional, and their clinical trial phases are listed in Figure 4.

**Figure 4:**
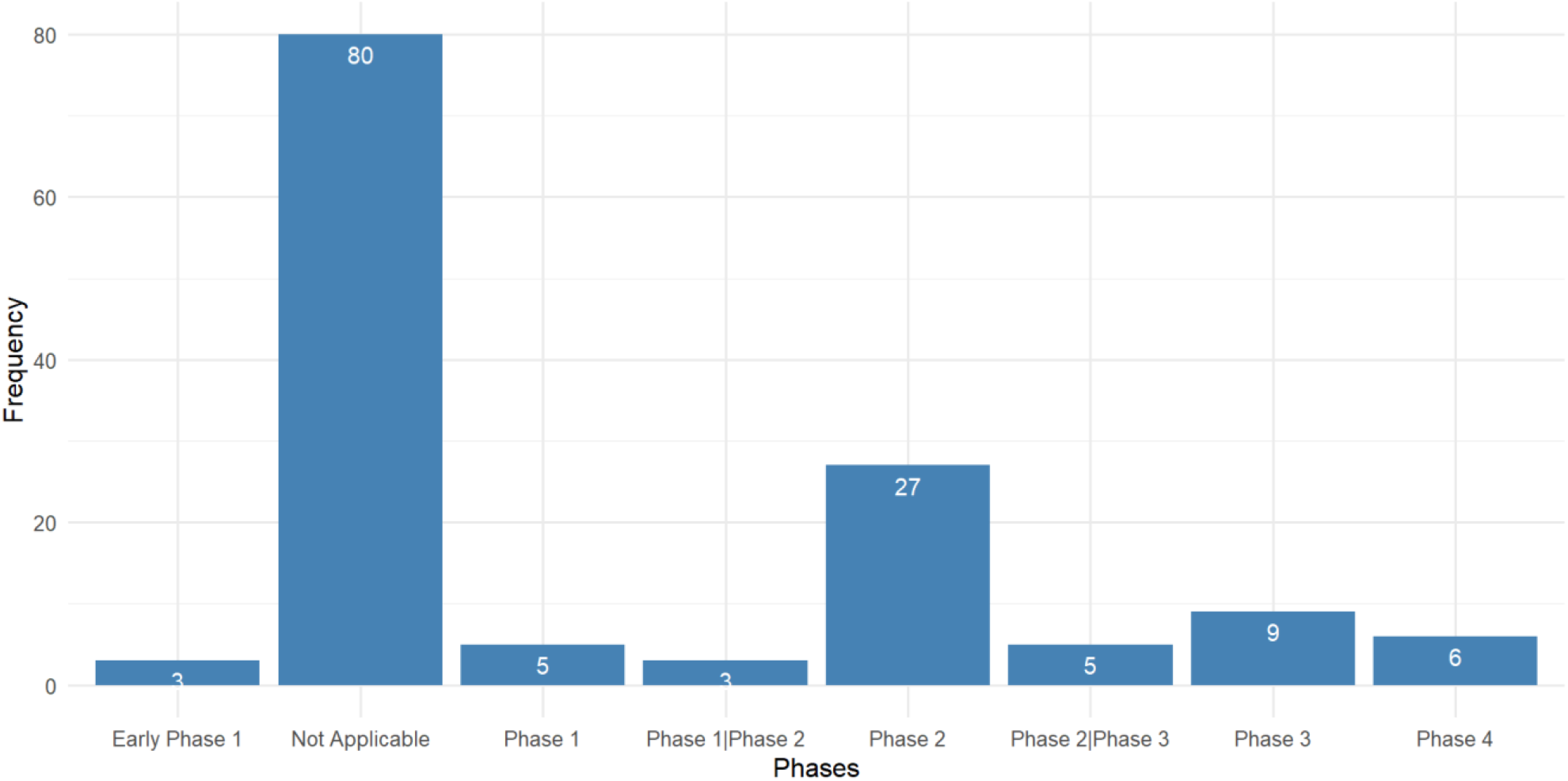
Study Type of Interventional Clinical Trials for Long- Covid-19

### Interventional Trials

The age group in the interventional trials have ∼48% with 18 Years and older inclusion criteria. The rest of them have some restrictions on the upper limit of age. Figure 5 shows the no. of participants enrolled in interventional trials by NCT IDs with NCT05449418 enrolling the largest number of 5000 participants followed by

**Figure 5:**
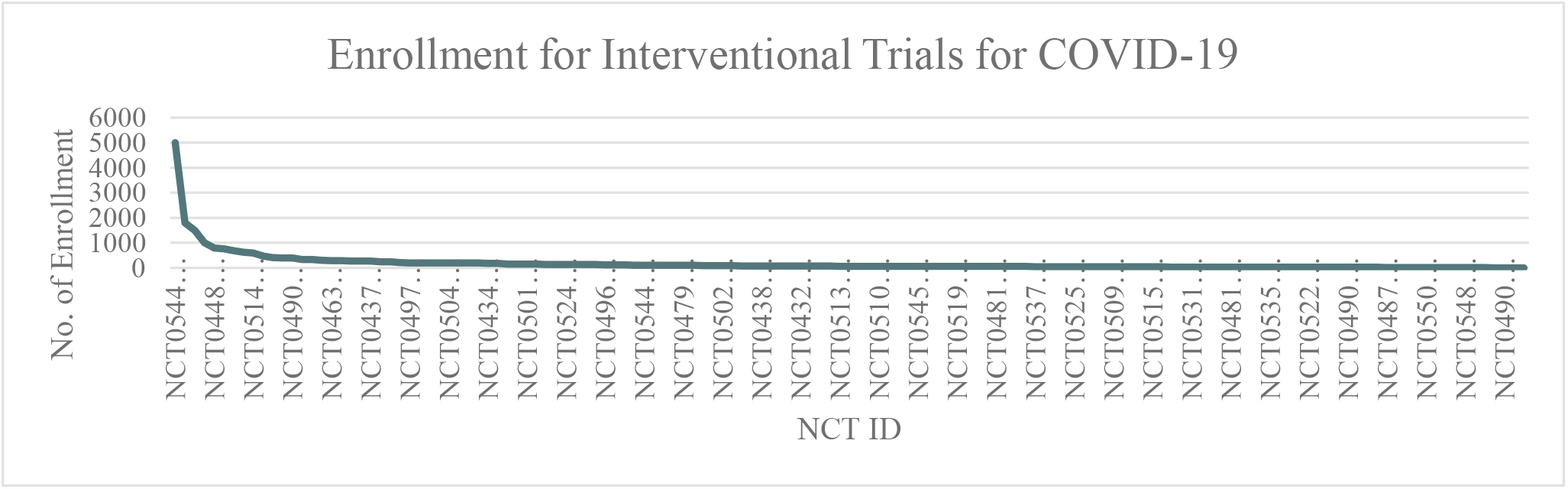
The no. of participants enrolled in Interventional Trials

NCT05168800, NCT04950725, NCT05513560 and NCT04481633. The details of the trials are present in A2. Figure 6 shows the total number of participants enrolled in the interventional trials by phases, where the total number of enrollment is maximum for phases that do not fall in any clinical trial phases.

**Figure 6:**
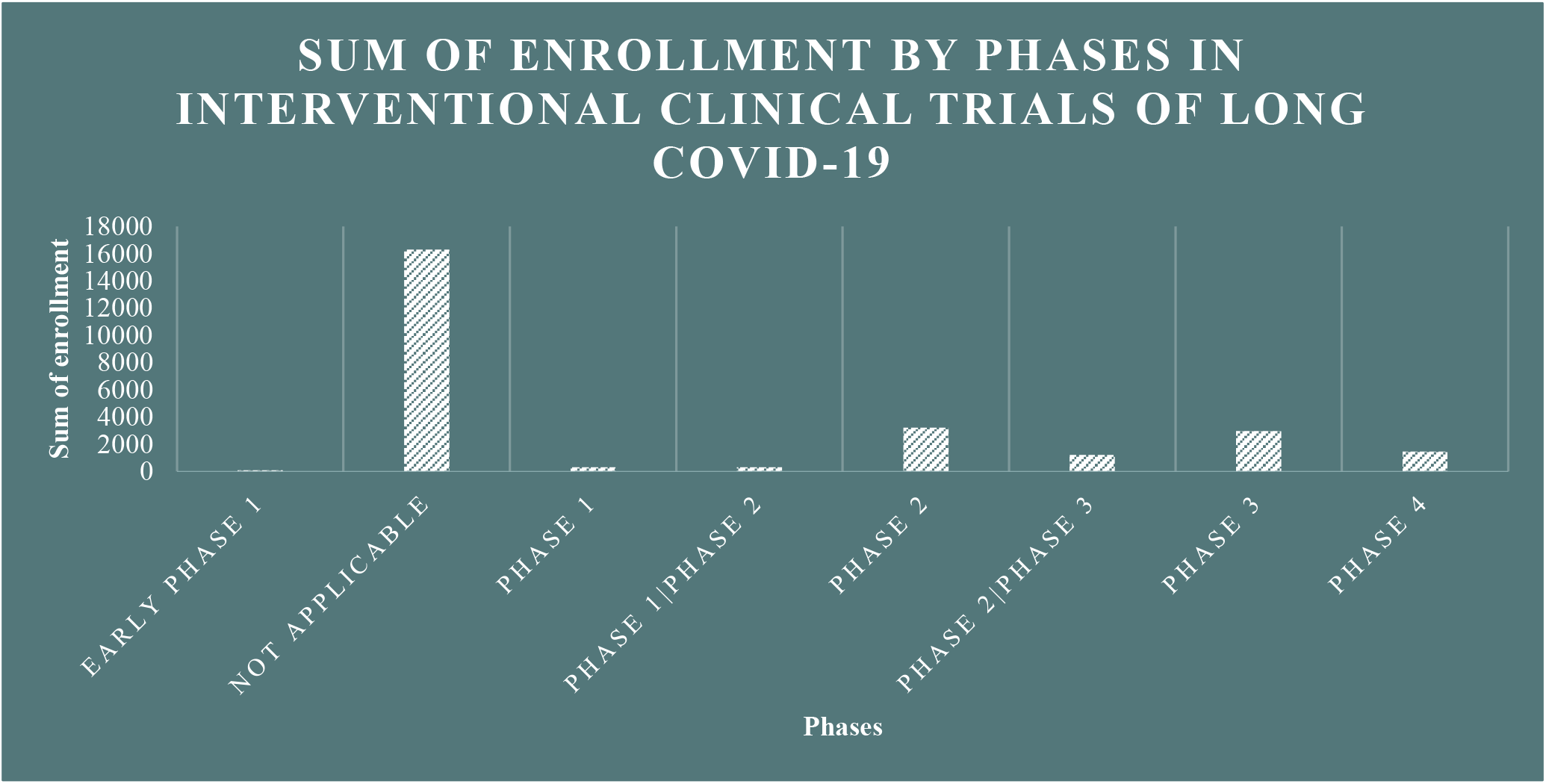
The sum of participants enrolled in Interventional Trials by Clinical Trial Phases

The time to completion of both types of trials (observational and interventional) is given in Figure 7 & 8. The maximum time length is 5 years for interventional trials

**Figure 7:**
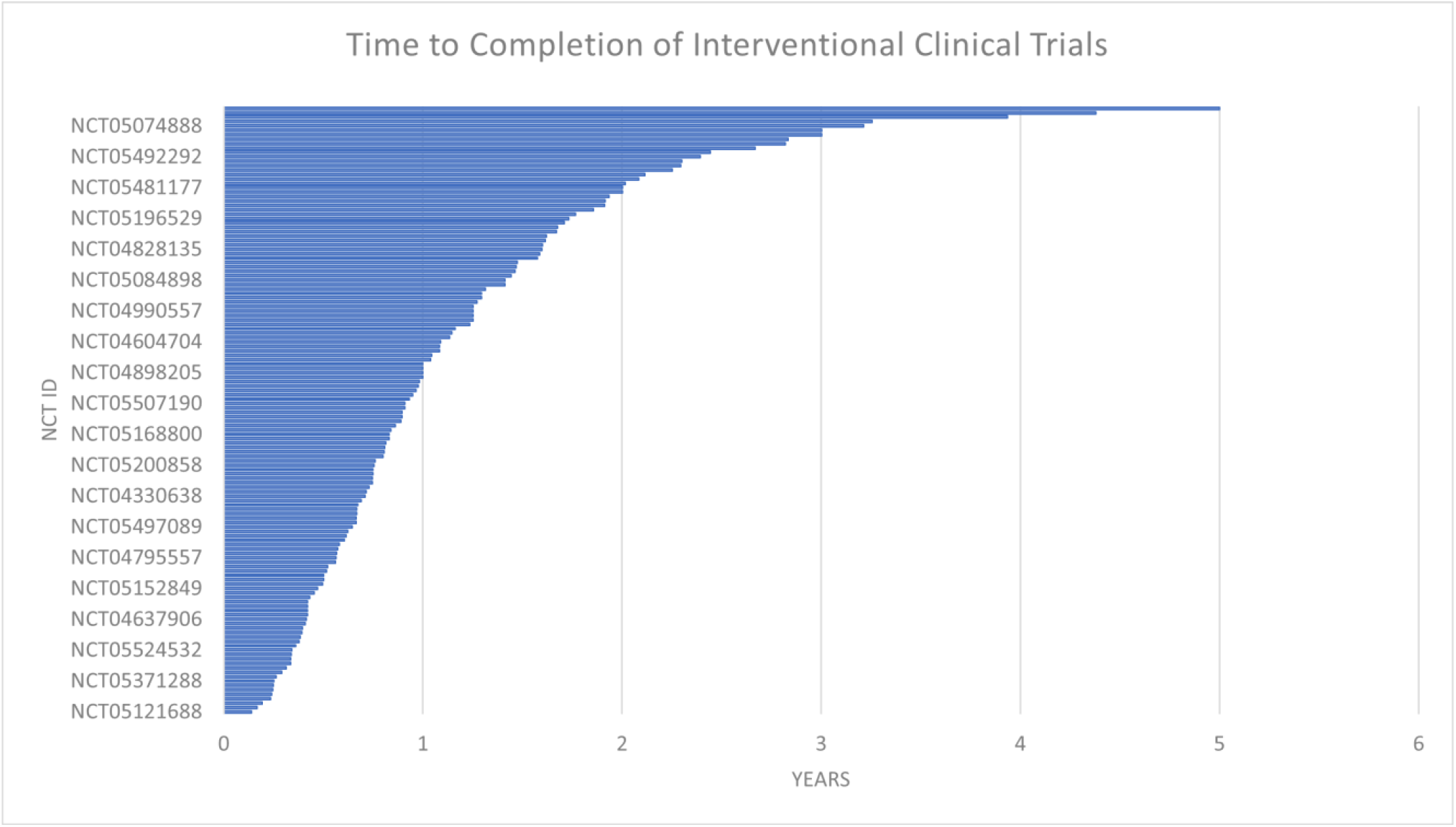
The time to completion of interventional clinical trials

**Figure 8:**
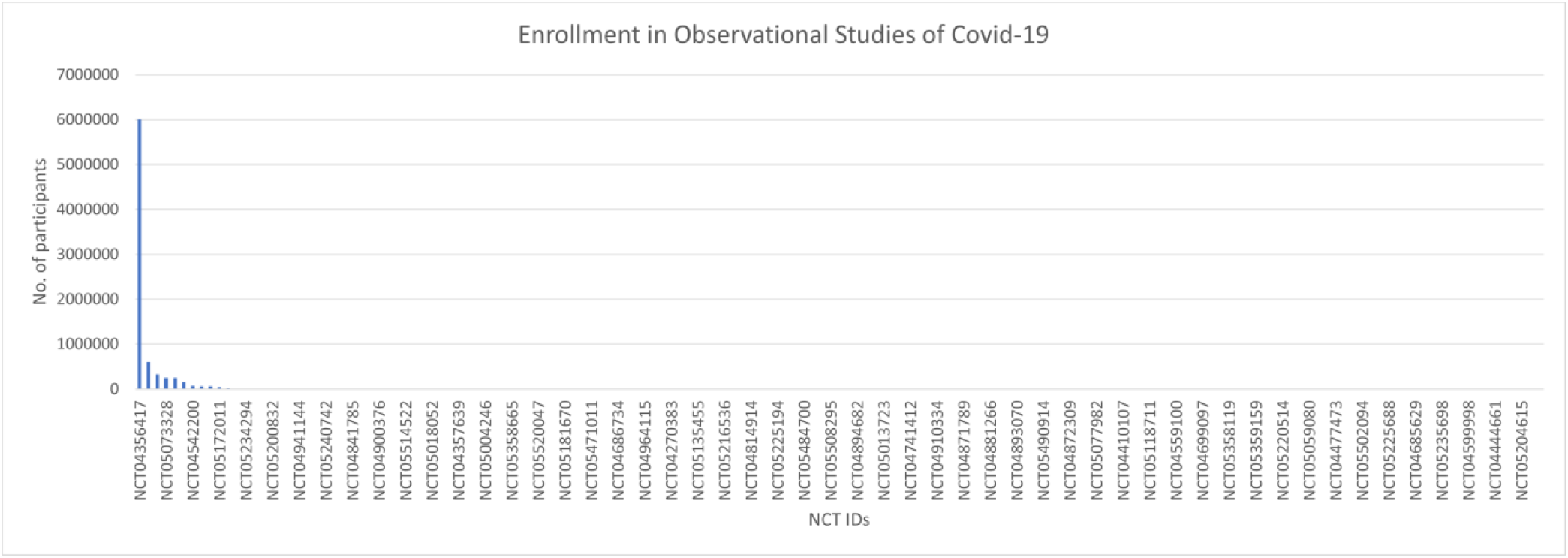
Enrollment in Observational Studies of Covid-19

**Figure 9:**
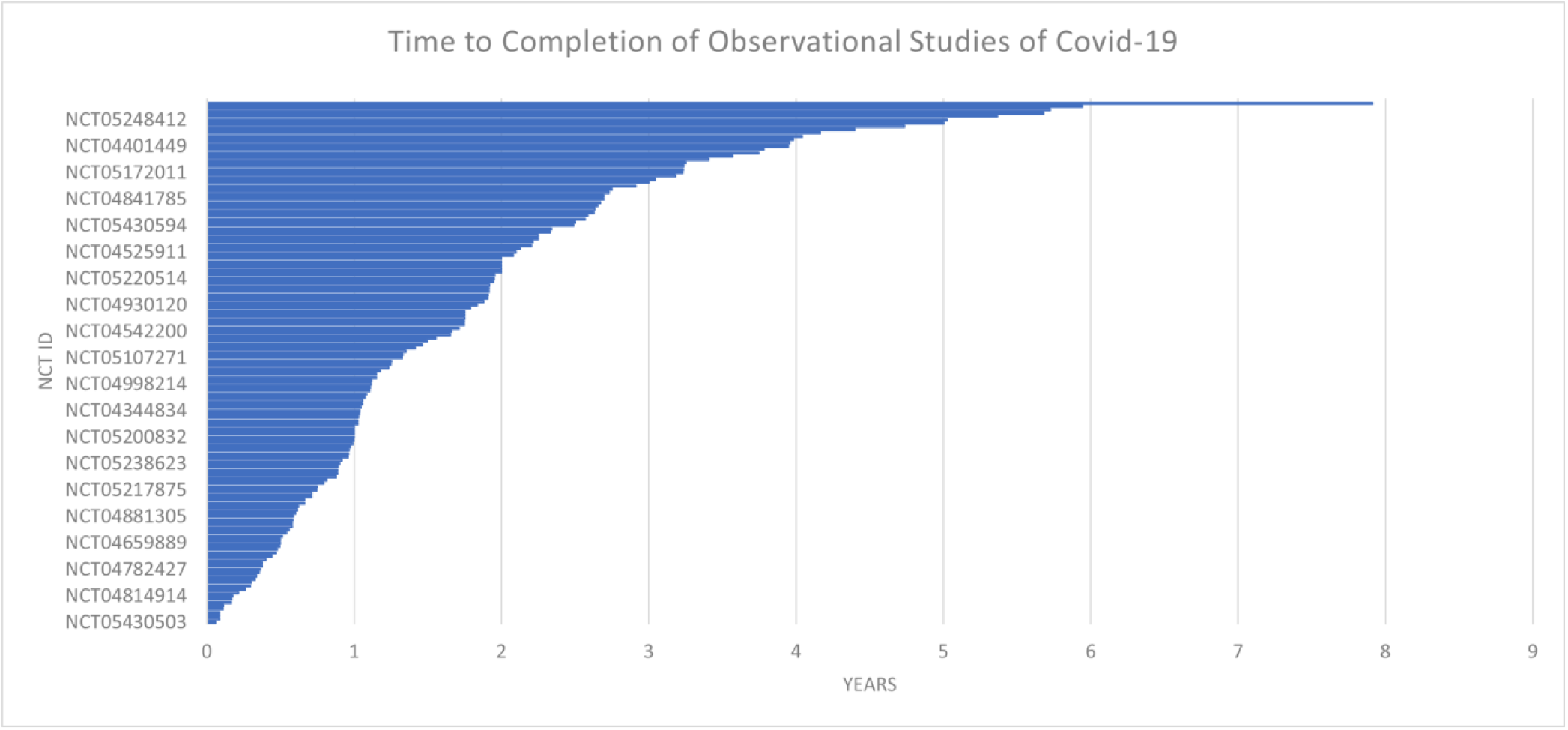
Time to Completion of Observational Studies of Covid-19 Pediatric Trials

### Observational Studies

There are 158 observational studies of which 1 is restricted to enrollment of males. The time for completion is a maximum of ∼8 years for an observational trial. The enrollment in observational trials is quite large and the maximum is NCT ID NCT04356417 with no. of participants of 6000000 There are four interventional trials that include children which are listed in table A1, and the trial numbers are NCT05373043 (funded by US Federal Government), NCT05445531, NCT04900961, and NCT05084898. The average enrollment is 207.75 participants with no restrictions on age. There are blind as well as open-label trials, with mostly parallel design. There are 30 observational studies that will include children.

## Analysis of Tweets from Social Media

We conducted a sensitivity analysis with 8346 tweets generated obtained from social media platform Twitter and wanted to see the current status of long -covid-19 and people ‘s opinions about it on social media. As we can see that there is a significant amount of both positive and negative sentiments in Figure 10. From figure 11, the word “longcovid” having the highest frequency, followed by “covid”. The word cloud gives a better picture in Figure 12.

**Figure 10:**
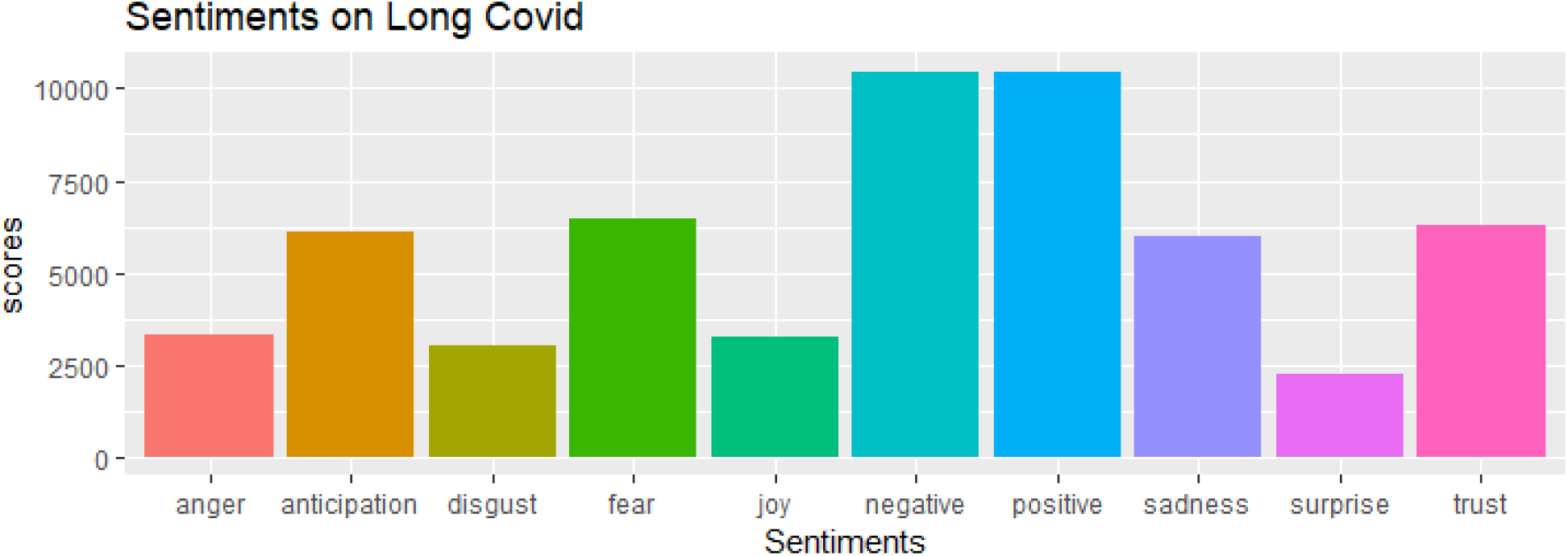
Sentiment Analysis of Tweets

**Figure 11:**
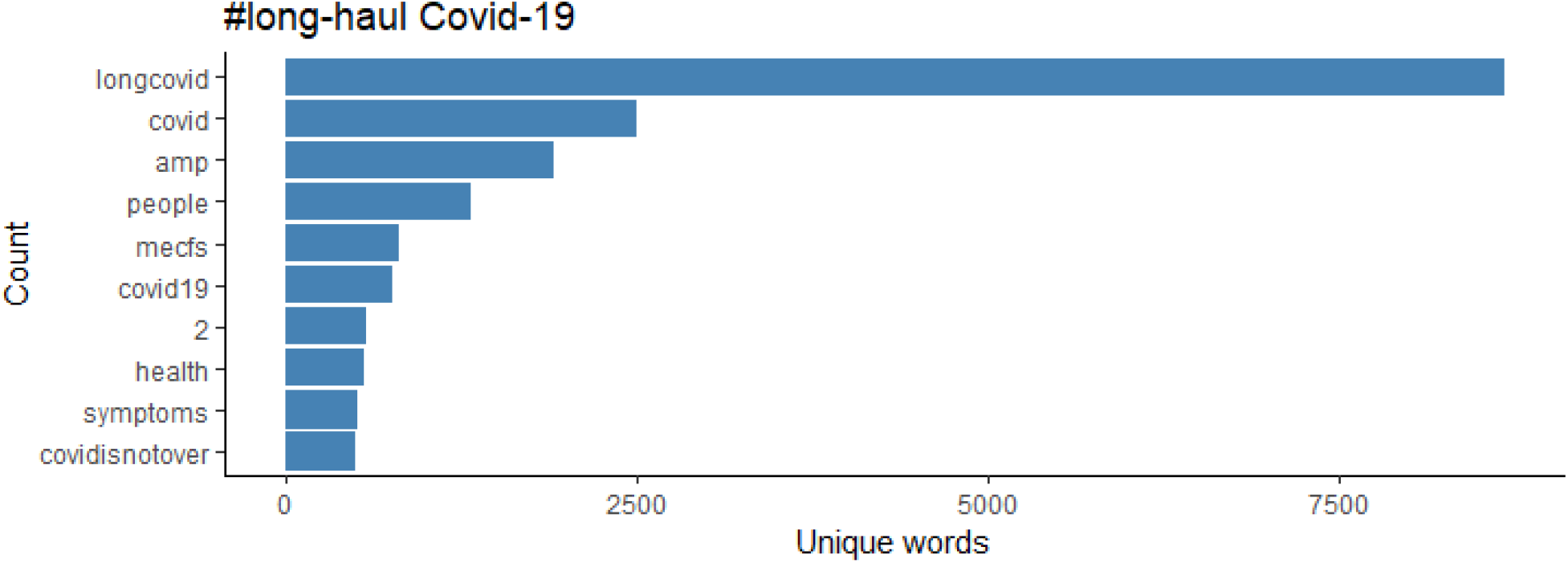
The top 10 words with the highest frequency

**Figure 11:**
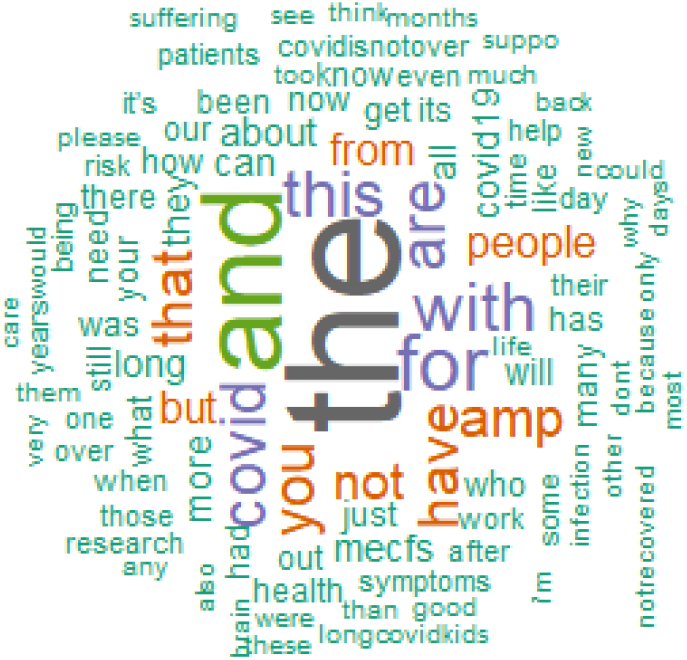
Wordcloud of the Tweets

**Figure 12:**
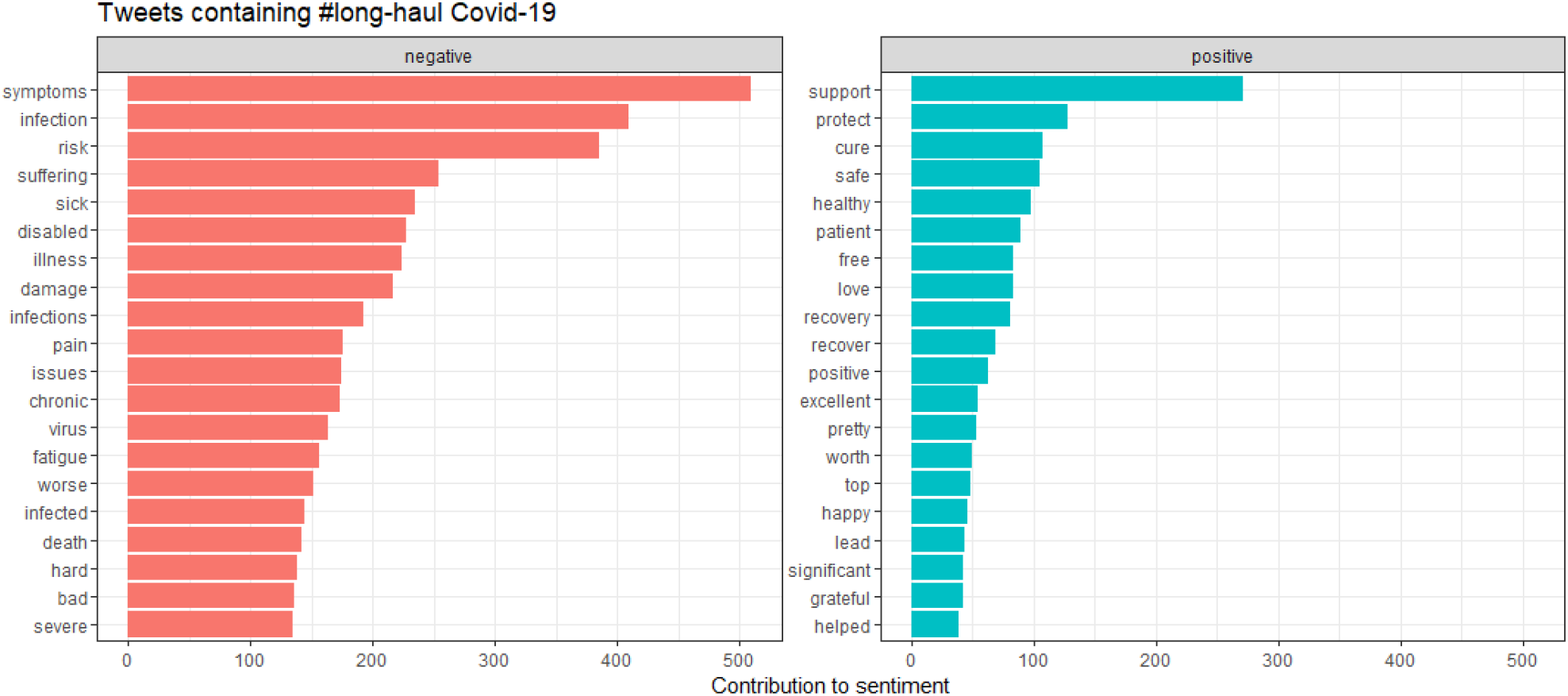
The contribution of positive and negative sentiments.

Figure 12 shows the contribution of words toward positive and negative symptoms, with Figure 13 showing the distribution of scores.

**Figure 13:**
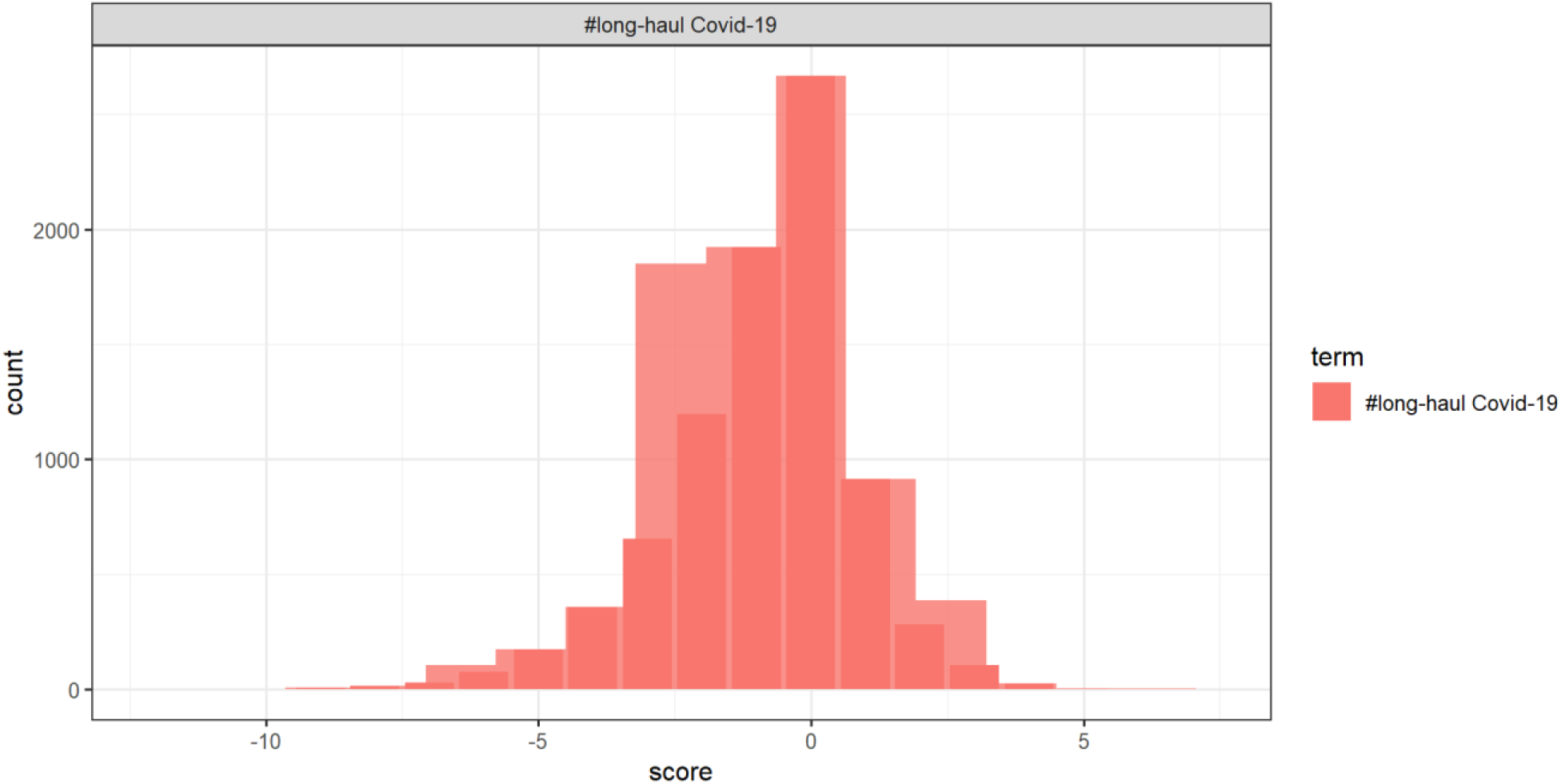
The distribution of sentiment scores.

**Figure 13:**
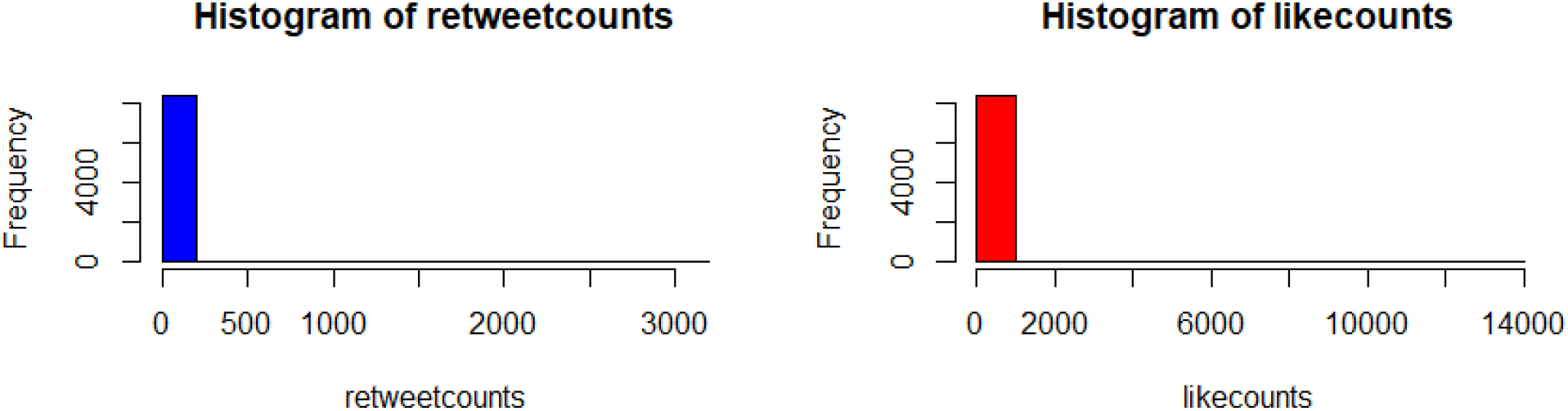
The histogram of the tweets shows an abundance of zeros

### Analysis of Tweet Sentiments by Negative Binomial Regression Model

After obtaining the sentiment scores for each of the observations through the “NRC” method of sentiment analysis, we wanted to test the hypotheses

1. The retweet counts are dependent on the sentiments
2. The favorite (like) counts are dependent on the sentiments.

We achieve these through Poisson, negative binomial and zero-inflated models. Count data regression modeling has received much attention in several science fields in which the Poisson, Negative binomial, and Zero-Inflated models are some of the primary regression techniques. Negative binomial regression is applied to modeling count variables, usually when they are over-dispersed. A Poisson distribution is used when the mean is equal to the variance. This situation is often unrealistic. The distribution of counts will usually have a variance that’s not equal to its mean. Modeling it as Poisson distributed leads to ignoring under- or overdispersion, depending on if the variance is smaller or larger than the mean. Also, situations with outcomes having a larger number of zeros require special needs of Zero-inflated models.

The histogram of the retweet counts and favorite counts are in Figure xx. The histogram shows a high proportion of zeros in the outcome measures. The covariates are “anger”, “anticipation”, “disgust”, “fear”, “joy”, “sadness”, “surprise”, “trust”, “negative”, and “positive”. The results by fitting all the count models are in Table 1 with outcomes “retweet counts”, and “favorite(like)counts”. The detailes estimates and p-values are listed in Table 2.

**Table 1:** Variable selection in models of retweet and favorite counts with sentiment scores

| Models | Poisson | Negative Binomial | Zero-Inflated (binomial logit link) | Comments |
| --- | --- | --- | --- | --- |
| Variables found Significant for retweetcounts | “anger”,<br>“anticipation”,<br>“disgust”, “fear”,<br>“joy”, “sadness”,<br>“surprise”, “trust”,<br>“negative”, “positive” | “disgust”, “joy”,<br>“sadness”, “surprise”,<br>“trust”, “negative”,<br>“positive” | “anticipation”, “joy”,<br>“sadness”, “trust”,<br>“positive” | Negative binomial model seems to be the best performing model. |
| Variables found Significant for favoritecounts | “anger”,<br>“anticipation”,<br>“disgust”, “fear”,<br>“joy”, “sadness”,<br>“surprise”, “trust”,<br>“negative”, “positive” | “anger”,<br>“anticipation”,<br>“disgust”, “fear”, “joy”,<br>“sadness”, “surprise”,<br>“trust”, “negative”,<br>“positive” | “positive” | Negative binomial model seems to be the best performing model. |

**Table 2:** Estimates (confidence intervals) from Models

| Variables | Negative Binomial Model with retweet counts as Outcome |  |  |  | Negative Binomial Model with favorite counts as Outcome |  |  |  |
| --- | --- | --- | --- | --- | --- | --- | --- | --- |
|  | Estimate | 2.50% | 97.50% | P-value | Estimate | 2.50% | 97.50% | P-value |
| (Intercept) | 5.189095 | 4.658966 | 5.793146 | 1.51E-218 | 20.61177 | 19.03433 | 22.35081 | 0 |
| anger | 1.102365 | 0.970846 | 1.253846 | 0.141028043 | 1.162999 | 1.053729 | 1.284773 | 0.002478 |
| anticipation | 1.065537 | 0.985909 | 1.153526 | 0.149869222 | 1.102797 | 1.039483 | 1.171189 | 0.003221 |
| disgust | 0.728357 | 0.632539 | 0.840187 | 4.04E-06 | 0.838794 | 0.754632 | 0.933522 | 0.000688 |
| fear | 0.921362 | 0.838073 | 1.013359 | 0.109780306 | 0.916722 | 0.854627 | 0.983612 | 0.024308 |
| joy | 0.81454 | 0.709756 | 0.935488 | 0.002939532 | 0.886361 | 0.801846 | 0.980654 | 0.020245 |
| sadness | 1.212522 | 1.093228 | 1.345493 | 0.00056631 | 1.215866 | 1.125497 | 1.313882 | 3.53E-06 |
| surprise | 0.709527 | 0.614592 | 0.82223 | 4.02E-06 | 0.783444 | 0.702947 | 0.874976 | 1.30E-05 |
| trust | 1.109723 | 1.007908 | 1.223671 | 0.027799061 | 1.086922 | 1.01049 | 1.170196 | 0.019462 |
| negative | 1.181363 | 1.086161 | 1.287272 | 0.00017715 | 1.134959 | 1.068147 | 1.207219 | 0.000158 |
| positive | 1.140512 | 1.054933 | 1.234997 | 0.000229091 | 1.093359 | 1.032497 | 1.158825 | 0.000907 |

## Discussion

The study aims to describe the prevalence of Long-haul Covid-19 among the local (US regions), and extrapolate our estimates to the global population. Also, we plan to estimate and compare the predictors of long-haul Covid-19. The study shows an urgent need in assessing the long-covi-19 situation. For further work, continuous variables can be presented by summary statistics (such as mean, median, standard error, range, 95% CI, and correlations), and the categorical variables by frequency distributions (frequency counts, percentages, and 95% CI). Simple linear regression, logistic regression, and Cox univariable and multi-variables regression analyses can be performed data specific. Survival curves for overall survival probability can be generated using Kaplan-Meier curves. Bayesian variable selection and machine learning algorithms can detect the causes related to the outcome of interest. The Chi-square test can be used to understand the association between two factors. Results can be declared significant at a significance level of 5% and with credible Bayesian intervals.

The main focus should be on building an appropriate prediction model that can incorporate various features such as vital signs, demographics, and genetic factors to predict the possibility of long covid-19 occurrence, including covid-19-related death prediction during the early stage of the study. A major initiative has been undertaken with L3C data challenge(12). The models developed may have both frequentist and Bayesian counterparts to address the uncertainities in the prediction alogorithms. The model need to have flexibility so that it can be modified according to the targeted population and outcomes, thus identifying the best set of predictors.

We can also address the progression of the disease through survival models. The solution to this problem involves determining the stages of disease progression and predicting the outcome of COVID-19 patients. Despite the use of patient inclusion and exclusion criteria, the total patient population comprises clinically relevant distinct subgroups identifiable by demographics, clinical histories, and molecular and disease characteristics. The International Council for Harmonization Efficacy guideline (ICH E9) discusses subgroups so that potential subgroups can be pre-planned to investigate treatment heterogeneity. So through the identification of the factors involved in predicting the long term outcome of Covid-19, we will be able to identify the subgroup of population who are at risk of such outcomes. Then, targeted treatment and medical resource allocation can be carried out for patients in different stages. Further, we can identify the subgroup of patients that have similar outcomes and demographics, lifestyle, or risk factors.

This research discusses reshaping several methodologies and research ideas in a single platform. The work can help to address several unaddressed research questions, thus in turn will help implement such prediction methods to address issues of designs in live clinical trials. The novel techniques and methodologies will help improve efficiency and identify subgroups that benefit from personalized treatment. Thus, in turn, the hands-on software can be utilized by clinicians and researchers to recommend doses, sample size, and power, and address the shortcomings in current trials, thus contributing to potential approval of therapies, and vaccines for long term efficacy contributing to the revenue of the company as a whole. Each design has its pros and cons and assumptions that must be justified. Similarly, who is at the risk of long COVID and what the future entails for long COVID patients is essential to characterize and understand. We will deal with a sicker population in the coming years, and we need healthcare resources to combat the post-pandemic lingering post-acute sequelae of COVID and other healthcare needs. The study of long COVID or broadly COVID survivorship is one of the areas where statisticians have a great deal to contribute in the coming years(10), targeting a population of more than 100 million affected worldwide, with 44 million people in the US. According to CDC, the current estimates show that 13.3% of the people with covid-19 have lasting effects at one month or longer after infection 2.5% at three months or longer, based on self-reporting, and more than 30% at 6 months among patients who were hospitalized. This shows that there is a potential market for drug development in a population with unmet research needs. The health effects of COVID-19 appear to be prolonged and can exert marked stress on the healthcare system. To understand the health care needs of the long-haul Covid-19 vulnerable population, research needs, dedicated clinics, and segregation of hospitalization facility needs are imperative to help the millions recovering from this disease. This multidisciplinary research article will encourage conducting clinical trials and channelizing the efforts toward long-term Covid-19-related therapeutic interventions to mitigate the adverse physical and mental health effects among the patients [8].

## Data Availability

All data produced are available online at https://clinicaltrials.gov/

